# Initial Whole Genome Sequencing and Analysis of the Host Genetic Contribution to COVID-19 Severity and Susceptibility

**DOI:** 10.1101/2020.06.09.20126607

**Authors:** Fang Wang, Shujia Huang, Rongsui Gao, Yuwen Zhou, Changxiang Lai, Zhichao Li, Wenjie Xian, Xiaobo Qian, Zhiyu Li, Yushan Huang, Qiyuan Tang, Panhong Liu, Ruikun Chen, Rong Liu, Xuan Li, Xin Tong, Xuan Zhou, Yong Bai, Gang Duan, Tao Zhang, Xun Xu, Jian Wang, Huanming Yang, Siyang Liu, Qing He, Xin Jin, Lei Liu

**Affiliations:** The Third People’s Hospital of Shenzhen, National Clinical Research Center for Infectious Disease, The Second Affiliated Hospital of Southern University of Science and Technology, Shenzhen 518112, Guangdong, China; BGI-Shenzhen, Shenzhen 518083, Guangdong, China; School of Medicine, South China University of Technology, Guangzhou 510006, Guangdong, China; BGI Education Center, University of Chinese Academy of Sciences, Shenzhen 518083, Guangdong, China; Guangdong Provincial Key Laboratory of Genome Read and Write, BGI-Shenzhen, Shenzhen, 518120, China; James D. Watson Institute of Genome Science, 310008 Hangzhou, China

## Abstract

The COVID-19 pandemic has accounted for more than five million infections and hundreds of thousand deaths worldwide in the past six months. The patients demonstrate a great diversity in clinical and laboratory manifestations and disease severity. Nonetheless, little is known about the host genetic contribution to the observed inter-individual phenotypic variability. Here, we report the first host genetic study in China by deeply sequencing and analyzing 332 COVID-19 patients categorized by varying levels of severity from the Shenzhen Third People’s Hospital. Upon a total of 22.2 million genetic variants, we conducted both single-variant and gene-based association tests among five severity groups including asymptomatic, mild, moderate, severe and critical ill patients after the correction of potential confounding factors. The most significant gene locus associated with severity is located in *TMEM189-UBE2V1* involved in the IL-1 signaling pathway. The p.Val197Met missense variant that affects the stability of the TMPRSS2 protein displays a decreasing allele frequency among the severe patients compared to the mild and the general population. We also identified that the HLA-A*11:01, B*51:01 and C*14:02 alleles significantly predispose the worst outcome of the patients. This initial study of Chinese patients provides a comprehensive view of the genetic difference among the COVID-19 patient groups and highlighted genes and variants that may help guide targeted efforts in containing the outbreak. Limitations and advantages of the study were also reviewed to guide future international efforts on elucidating the genetic architecture of host-pathogen interaction for COVID-19 and other infectious and complex diseases.

## Introduction

It has been more than 100 years since the 1918 influenza outbreak killed at least fifty million people worldwide^1^. Now we are facing another pandemic. Since the late December of 2019, the 2019 novel coronavirus diseases (COVID-19) has spread rapidly throughout the world, resulting in more than five million confirmed cases and hundreds of thousands deaths in less than six months^2,3^. The disease was caused by the infection of a novel enveloped RNA betacoronavirus that has been named severe acute respiratory syndrome coronavirus 2 (SARS-CoV-2), which is the seventh coronavirus species that causes respiratory disease in humans^4,5^. The virus causes serious respiratory illnesses such as pneumonia, lung failure and even death^6^. Until now, there is no specific therapeutics and vaccine available for its control. Continuing epidemiological and molecular biological study to better understand, treat and prevent COVID-19 are urgently needed.

A characteristic feature of many human infections is that only a proportion of exposed individuals develop clinical disease and for the infected persons, severity varies from person to person^7^. In the COVID-19 outbreak, a high level of inter-individual variability was observed in terms of disease severity and symptomatic presentation. Around 80%-85% of the laboratory confirmed patients were classified as mild (i.e. nonpneumonia and mild pneumonia) while 15%-20% would progress to severe or critical stage with a high probability of respiratory failure^8–11^. Patients with severe disease had more prominent laboratory abnormalities including lymphocytopenia and leukopenia than those with non-severe disease^12,13^. In addition, not all people exposed to SARS-CoV-2 were infected according to the epidemiological observation of the patients’ close contacts^14,15^. Notably, previous studies have indicated that genetic background plays an essential role in determining the host responses to infections by HIV^16–18^, HBV^19^, HCV^20^, influenza^21–24^, SARS-CoV^25,26^ and numerous common viruses^27^ etc. Those studies highlighted the HLA alleles and several genes involved in the interferon production and viral replication pathway and indicates that genetic factors may also play an important role to explain the inter-individual clinical variability among patients infected by SARS-CoV-2.

Till now, the global genetic community has been actively investigating in the genetic contribution to COVID-19. A recent twin study in UK suggests a 30% −50% genetic heritability for self-reported symptoms of COVID-19 and the predictive disease onset^28^, indicating a very strong genetic background predisposing the COVID-19 patients’ clinical manifestation and susceptibility. An earlier studies comparing the distribution of ABO blood group from 1,775 patients infected with SARS-CoV-2 with 3,694 normal people from Wuhan city and 23, 386 people from Shenzhen city suggested that blood group A had a significantly higher risk for COVID-19 (OR=1.20, p=0.02) while blood group O had the lower risk^29^. Using allele frequency and expression quantitative loci (eQTL) information of general healthy population from 1000 genome project and others, a few studies investigate the mutation frequency spectrum in different populations in candidate genes such as *ACE2* and *TRMPSS2*^30–32^. Genome-wide association test on array data from the UK Biobank participants with a positive and negative PCR-tests also reveals a few suggestive genes^27^. The COVID-19 host genetics initiative was established to encourage generation, sharing and meta-analysis of the genome-wide association summary statistics data around the world^33^. International collaborative efforts are necessary to elucidate the role of host genetic factors defining the severity and susceptibility of the SARS-CoV-2 virus pandemic.

Herein, we report the first genetic study of COVID-19 disease severity in China by deeply analyzing the association between the genetic variants present in the patients’ genome and their disease progression. We have recruited 332 hospitalized patients from a designated infectious disease hospital in Shenzhen City^34^. The patients display varying clinical and laboratory features and were categorized as asymptomatic, mild, moderate, severe and critical cases according to the criteria made by the Chinese Center for Disease Control and Prevention^6^. To maximize the statistical power given the relatively small hospitalized sample size and for accurate detection of extremely rare variants, we conducted deep whole genome sequencing (average 46x) for the patients. Given a fixed samples size, this protocol facilitates the estimation of genetic effects of rare and loss of function variants in addition to the common variants that may be potentially contributing to the COVID-19 clinical variability^35^. Based on the 22.2 million variation detected from the patients, we investigated host factors by conducting both single variant and gene-based genome-wide association study and by evaluating the difference of allele frequency of the protein truncating variants and HLA alleles among the patient groups. In addition, we performed joint-calling of the genetic variants of the unrelated COVID-19 patients (N=284) and the publicly available Chinese genomes from the 1000 genome project^36^ (N=301, ∼7x) and 665 selected Chinese genomes from the Chinese Reference Panel Population (manuscript in preparation, ∼30x) to explore potential genetic factors that may contribute genetic susceptibility of SARS-CoV-2 infection.

## Results

### Clinical and laboratory features of the 332 hospitalized COVID-19 patients

The 332 recruited patients with laboratory-confirmation of SARS-COV-2 infection were being quarantined and treated in the Shenzhen Third Hospital. We extracted and analyzed the clinical symptoms, laboratory assessment, recent exposure history of the patients from the hospital’s electronic medical records. The 332 patients consist of 48 family members and 284 unrelated individuals.

25 (7.5%), 12 (3.6%), 225 (67.8%), 53 (16.0%) and 17 (5.1%) patients were defined as asymptomatic, mild, moderate, severe and critically severe according to the most severe stage they encountered during the disease course following the Chinese CDC criteria^6^ **(Figure 1A)**. The asymptomatic, mild and the moderate groups of patients had positive RT-PCR test result but did not have or only had mild pneumonia. The severe patients met any one of the following criteria: respiratory rate (RR) ≥ 30/min, blood oxygen saturation ≤ 93%, partial pressure of arterial oxygen to fraction of inspired oxygen ratio (PaO2/FiO2) < 300 mmHg and/or lung infiltrates > 50% within 24-48 hours. A severe patient was classified as critical ill if he/she experienced any one of the following situations: respiratory failure, septic shock and/or multiple organ dysfunction or failure. A broader definition of the mild group includes the asymptomatic, mild and moderate patients, and of the severe group, includes the severe and critically severe patients.

**Figure 1.**
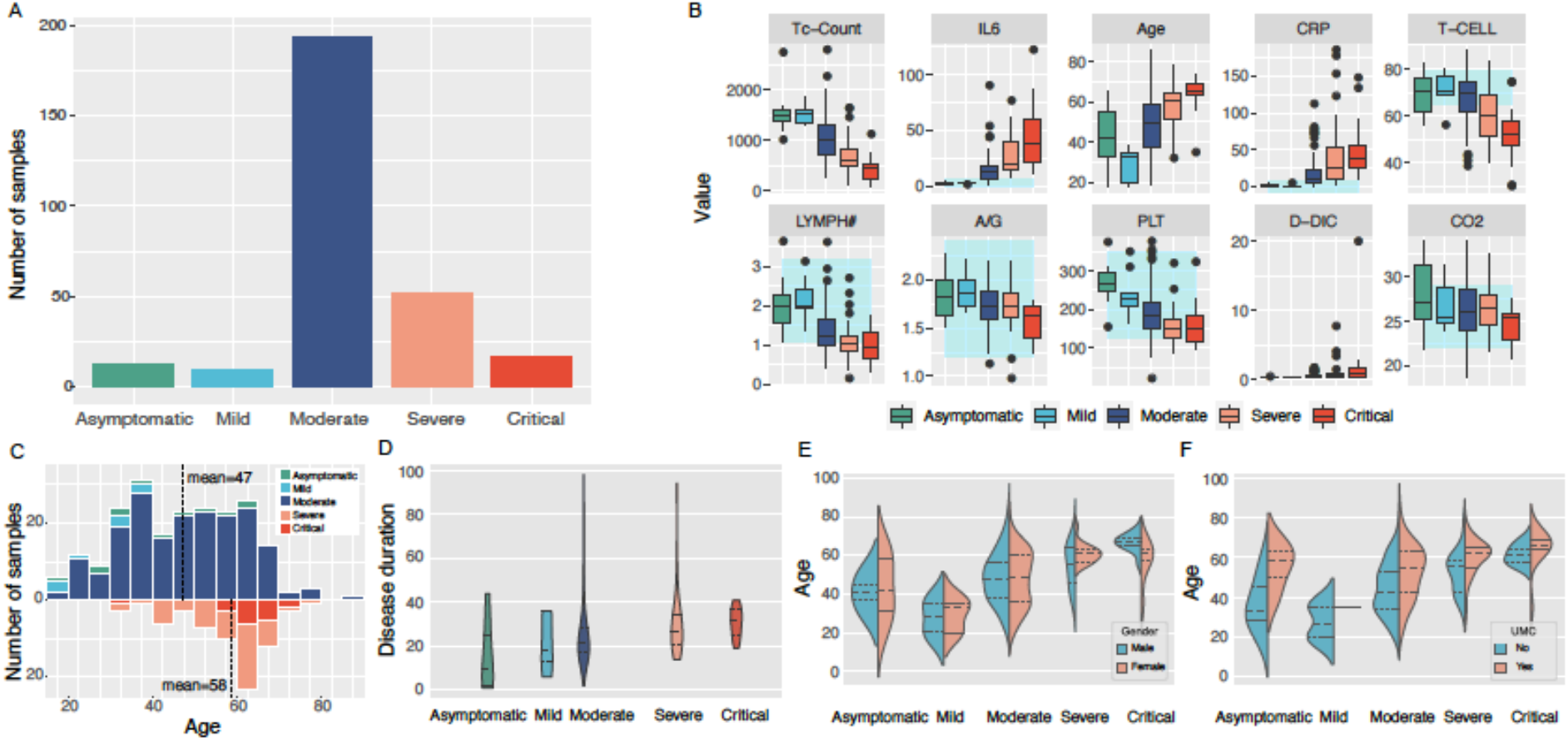
Clinical and laboratory assessments of the recruited 332 COVID-19 patients. (A) number of samples belong to the five categories (B) top 20 features that classify the patient categories in the machine learning trained model (C) age distribution for the five categories of patients (D) distribution of disease duration, i.e. the duration between the disease onset and the first negative RT-PCR test among the five groups of patients (E) gender distribution for the five categories of patients by age (F) distribution of the proportion of patients with or without medical comorbidities among the five categories of patients by age.

The patients displayed several clinical presentations typical to COVID-19, which mainly involved fever (70.8%), cough (54.2%), fatigue (23.9%), hoarse voice (17.6%), loss of appetite (16.2%), delirium (15.1%) **(Figure S1)**. Less than 10% had also experienced diarrhea, chest and abdominal pain, shortness of breath and anosmia. More than 50% of the patients had at least one medical comorbidities (e.g., hypertension). Consistent with previous report, the broadly defined severe patients tend to be older (severe average 45 years old vs mild average 58 years old, t-test p=0.03, **Figure 1C**), suffer from a longer course of disease between the onset and the first negative RT-PCR test outcome (**Figure 1D**) and shorter exposure time (**Figure S2**). In addition, the severe patient group consist of more males than females (severe 66.7% vs mild 41.3%, χ^2^ test p=4.3e-4, **Figure 1E**) and tend to undergo medical comorbidities more frequently (severe 58.8% vs mild 45.1%, χ^2^ test p=0.07) (**Figure 1F**) than the mild patients.

During hospitalization, a series of sixty four laboratory assessments including a complete blood count and blood chemical analysis, assessment of liver function, assessment of renal functions, test of humoral immunity, test of coagulation, measure of electrolyte and measure of blood gas electrolyte (**Figure S3**) and a time-series evaluation of T lymphocyte subgroups (**Figure S4**) were performed for each of the patients to monitor their disease status and progression. Using a tree-based machine learning prediction model^37^, we computed the local interaction effects of the sixty four laboratory assessment features as well as three demographic features including age, gender and w/o medical comorbidities for classification of the patient severity category (**Figure S5**). The top ten features of greatest importance that contribute to a severer disease outcome include decreased lymphocyte counts (Tc-Count, T-CELL, LYMPH#) and platelet counts, evaluated interleukin 6, C-reactive protein and D-dimer, increased age and decreased A/G and CO2 (**Figure 1B**), consistent with previous reports^38^. We applied the top twenty features of importance to assign a severity score for each patient to reflect their disease status (**Figure S6**).

### Deep whole genome sequencing and genetic variation

We obtained the whole blood and performed deep whole genome sequencing for the recruited patients. There is no significant difference for sequencing depth between the broadly defined mild and severe group (mild 46.26x vs severe 46.71x) (**Figure 2A**). We conducted variation detection and genotyping using the GATK joint genotyping framework to avoid any potential batch effect derived from individual variant calling. Bioinformatics analysis and the data quality control process were described in details in the **Online methods**.

**Figure 2.**
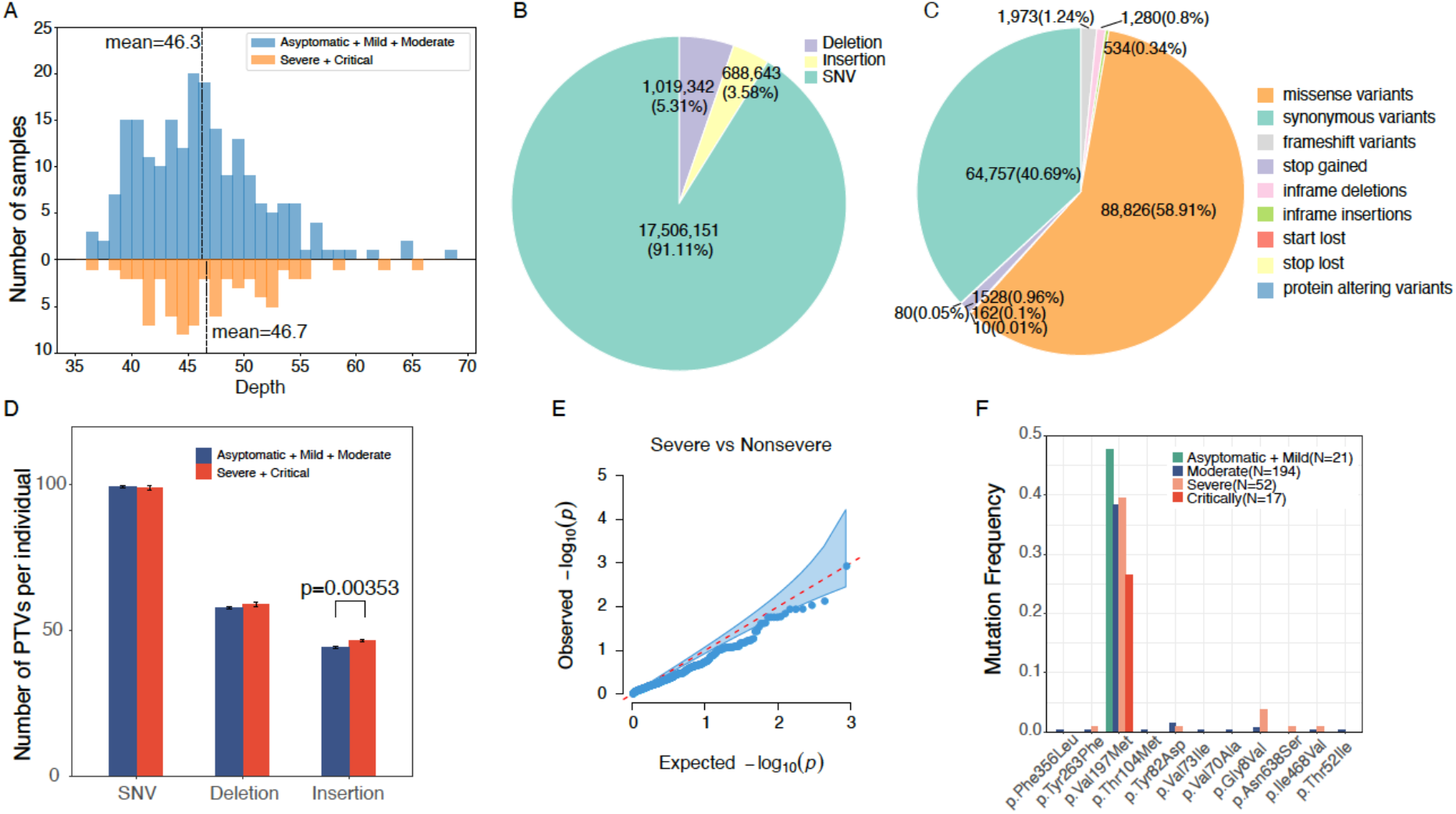
Deep whole genome sequencing and genetic variation among the patients. A) Sequencing depth distribution B) Proportions and numbers of types (SNP, Indel) of genetic variants identified from the patients C) Proportions and numbers of functional consequences of the genetic variants among the patients D) comparison of loss of function variation burden for SNP, small insertions and deletions between the severe and the non-severe patients E) Single variation association test for loss of function mutation burden between the severe and non-severe patients F) allele frequency distribution for all the missense and loss of function variants present in *ACE2* and *TRMPSS2* genes.

Among the 332 patients, we identified a total of 22.2 million variants including 17.9 million bi-allelic single nucleotide polymorphism, 1.75 million bi-allelic small insertions and deletions and 2.49 million multi-allelic variants **(Figure 2B)**. The average transition/transversion (ts/tv) ratio is 2.12 and the proportion of heterozygous versus homozygous variants among all the samples is 1.29, consistent with our expectation^39^ and indicates good quality of the variant calls (**Figure S7**). Particularly, we have identified 398K variants that result in an alteration of the protein coding sequence (**Figure 2C**). The QC metrics were detailed in **Table S1**.

Our first question was whether the most vulnerable severe and critical patients may have a monogenic basis for their demonstration. We investigated the burden of loss of function variants predicted by the ensemble variant effect predictor among the patient groups^40^. In total, we have identified 4,891 predicted loss of function variants including 1,860 frameshift, 1,447 stop gained, 505 splice donor and 380 splice acceptor variants among the 332 patients. On average, each patient possessed 201 predicted loss of function variants in their genome **(Figure S8)**. 261 of those variants were uniquely presented in the COVID-19 patients (18.6%) and have not been previously reported in the 1000 genome and the gnomAD studies^36,41,42^. Interestingly, the severe and the critical patients tend to have more loss of function insertions than the asymptomatic, mild and the moderate groups in a logistic regression taking the number of loss of function variants as variable and the patients’ age, gender, the twenty principle components and effective sequencing depth as covariates (p=0.004) (**Figure 2D**). When performing a mutation burden test for each of the 16,801 genes that have more than one variant among the 284 unrelated patients, we didn’t identify genes that were enriched in loss of function variants in the severe and critical patients (**Figure 2E**). On the other hand, we found two heterozygous loss of function variants located in *MST1R* and *RASA2* that were only present in the asymptomatic patients (**Figure S9**). The *MST1R* encodes the macrophage stimulating 1 receptor expressed on the ciliated epithelia of the mucociliary transport apparatus of the lung and follows an autosomal dominant inheritance mode for susceptibility to nasopharyngeal carcinoma^43^. Because those loss of function variants were only present in one patient, we didn’t build up links to the COVID-19 severity.

Particularly, we have inspected the missense and loss of function variants present in the SARS-CoV-2 S protein host cellular receptor gene *ACE2* and the S protein primer gene *TMPRSS2* that plays a critical role in controlling the viral entry into the host cell, as well as a few other genes that were predicted to play a role in the host pathogen interaction network like *SLC6A19, ADAM17, RPS6, HNRNPA1, SUMO1, NACA* and *BTF3* ^44^. The majority of the functional variants have minor allele frequency less than 1% except for the p.Val197Met missense variant in *TMPRSS2* **(Figure 2F**). Although not statistically significant, the p.Val197Met variant (rs12329760) displays a higher allele frequency in the asymptomatic and mild group compared to the rest of the group (asymptomatic: 0.46, mild: 0.50, moderate: 0.38, severe: 0.39, critical severe: 0.26). p.Val197Met was previously found to have higher allele frequency in East Asian (0.31-0.41) and Finnish (0.36) but is less frequently seen in South Asians (0.14-0.29) and the Europeans (0.17-0.23) (**Figure S10**). By computational protein modelling, the p.Val197Met TMPRSS2 isoform could decrease the stability of the TMPRSS2 protein, promote the binding to S-protein and inhibit its binding with ACE2^44^. The decreasing allele frequency in the severe patient groups supports that the p.Val197Met is related to the disease outcomes of COVID-19. The other genes didn’t contain significant allele frequency difference among the patient groups (**Figure S11**).

### Genetic association of common and rare variants with COVID-19 severity

To further investigate genetic effects for the patient severity, we performed genome-wide single variant association test and sequence kernel association test (SKAT) analysis of three traits implicating patient severity. We defined the first trait as a dichotomous classification of the broadly defined “severe group” that consists of the severe and critical ill patients (N=70) and the “mild group” (N=262) that consists of the asymptomatic, mild and moderate patients. We defined the second trait as a quantitative measurement of the severity level trained from the demographic features such as age, gender and the sixty-four laboratory assessments (N=332) (**Figure S5-6**). We used the disease duration from the electronic health records as the third trait which corresponds to the duration of time between the complained disease onset and the first laboratory confirmed PCR-test negative outcome (N=233) (**Figure 1D**). Power analysis indicates that given 80% statistical power, we will be able to identify associations between genotypes and phenotypes for variants with minor allele frequency greater than 0.2 and with a relative genetic risk contribution greater than 2 given the current sample size for dichotomous trait and similarly for the quantitative trait (**Figure S12**). Principal component analysis of the patients suggests little genetic differentiation (**Figure S13-14**).

We tested all the QC-passed 19.6 million bi-allelic variants for association with each of three traits in a logistic or linear regression model that includes gender, age, and the top 20 PC axes as covariates. The global distribution of resulting p-values was very close to the null expectation (λ = 0.996∼1.1, **Figure S15**) indicating that stratification was adequately controlled. The most significant SNP rs6020298 is located in the intron of a read-through transcript *TMEM189-UBE2V1* in the 20q13.13 region. (**Figure 3A-B**). The rs6020298 (hg38 chr20:50152518, A allele frequency severe vs non-severe: 0.59 vs 0.45) marks a suggestive significant association signal for both the Severe and Mild binary trait (logistic regression p=4.1e-6, OR=1.2) and the quantitative measurement of the severity score (linear regression p=1.1e-6, beta=0.35). SNPs in linkage disequilibrium with rs6020298 (r^2^>0.8) also affect the gene *UBE2V1* and *TMEM189* (**Figure 4A**). The *UBE2V1* gene encodes the ubiquitin-conjugating enzyme E2 variant 1. Both the UBE2V1 and TMEM189-UBE2V1 have been involved in the interleukin-1 (IL-1) signaling pathway^45^ and suggested to work together with TRIM5 to promotes innate immune signaling^46^. IL-1 is elevated in COVID-19 patients especially the severe and critical patients who suffer from the cytokine storm and severe inflation^47^. Clinical trial using IL-1 blockade on critical patients results in an improvement in respiratory function in 72% of the patients^48^. The lead SNP rs6020284 has a minor allele frequency close to 0.5 among the worldwide populations except for the African population (AF=0.13) (**Figure 4B**). It is also an eQTL for *LINC01273, TMEM189* among several tissues including the lung where the risk A allele increases the TMEM189 and LINC01273 expression in several tissues (**Figure S16**). This may indicate that an inborn evaluated TMEM189 expression in the patients may promote IL-1 signaling and predisposes the patients towards a poorer outcome against the COVID-19 infection. However, given the limited sample size in this study and that the intermediate pathways between TMEM189 and IL-1 production is still unclear, more replication and functional validation efforts should be made to re-evaluate this association signal. Notably, the TMEM189-UBE2V1 locus has been associated with monocyte percentage of leukocytes and granulocyte percentage of myeloid white cells^49^. Nonetheless, we didn’t observe nominal association (p<0.05) at the lead SNP rs6020298 with all the sixty-four laboratory assessments among the patients (**Figure 4C**). Therefore, the observed signal is not supposed to be confounded by individual variability on blood cell types. There is no strong genetic association with the disease durations **(Figure 3C)**.

**Figure 3.**
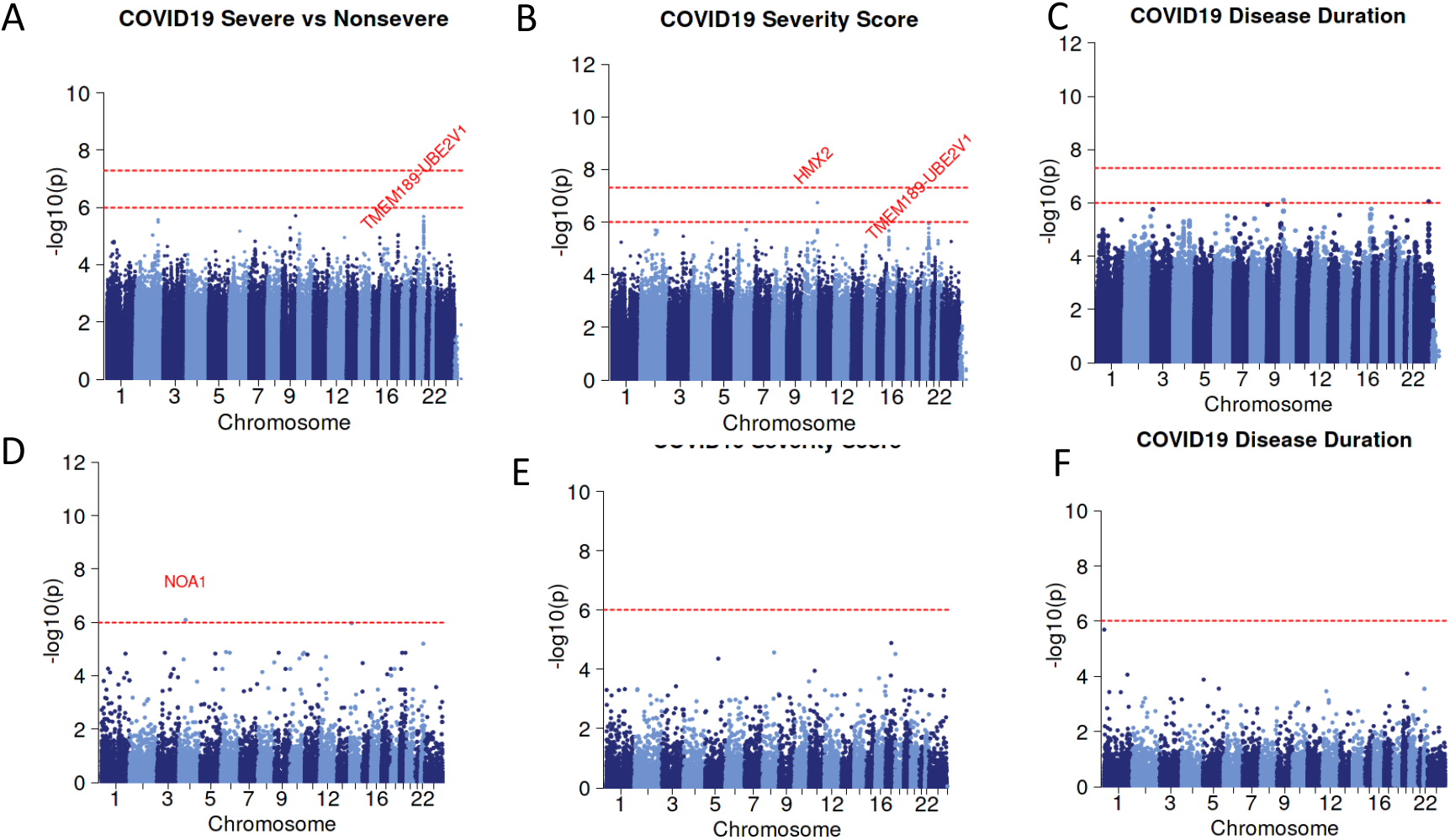
Genetic loci associated with patient severity. (A)-(C) Single variant and association test for three severity traits. (A) Severe and critical severe groups versus the rest of the non-severe groups. (B) Severity score assessed by laboratory test measurements. (C) the duration from disease onset to recovery (D)-(F) Gene-based association test for three traits.

**Figure 4.**
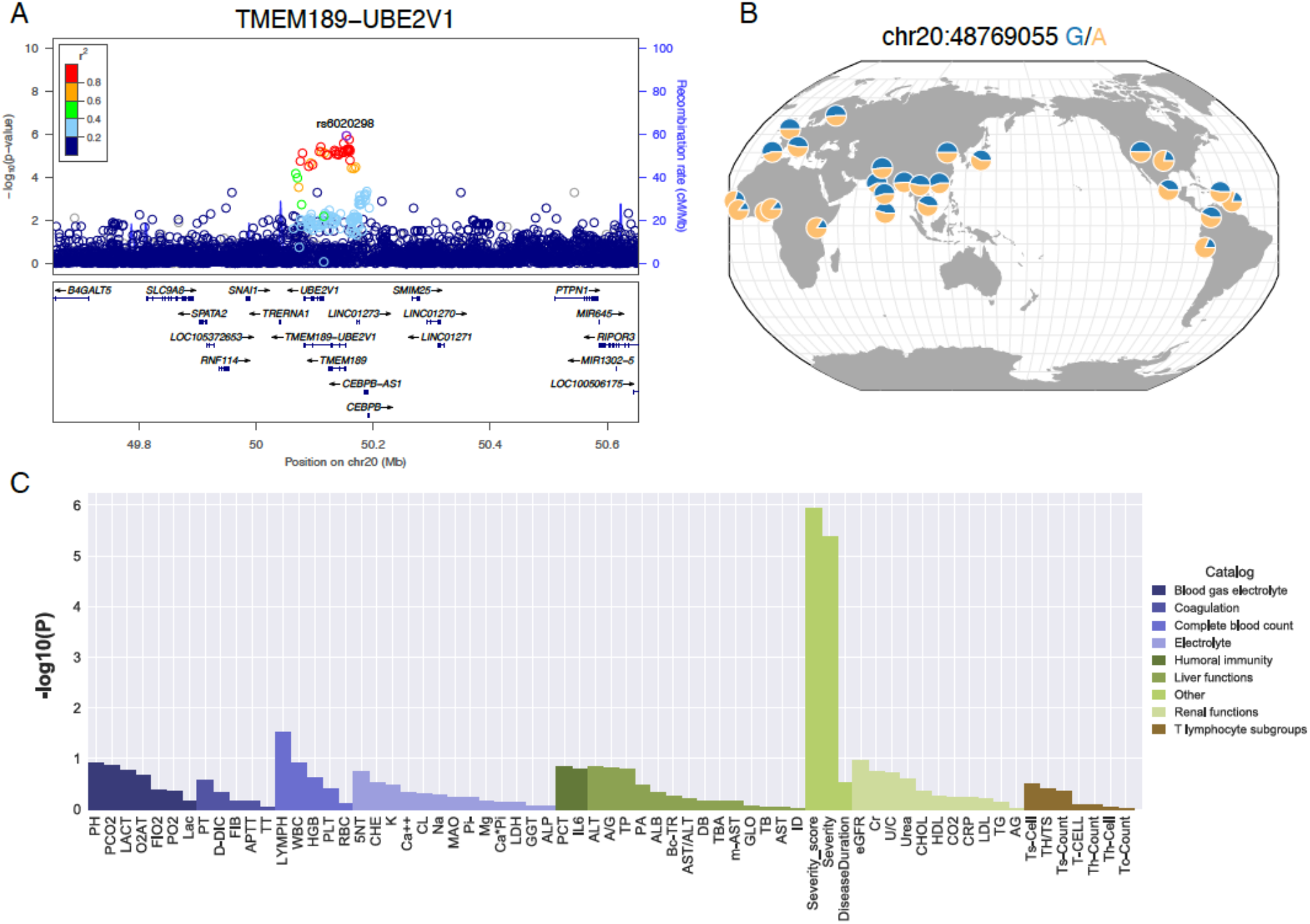
LD, allele frequency and pleiotropic effects of the *TMEM189-UBE2V1* signal suggestively associated with COVID-19 patient severity. A) Locuszoom plot shows the p-value of the SNPs centering the lead SNP rs6020298 and the recombination rate. Color of the dots indicate linkage disequilibrium r2 metric. B) Allele frequency of s6020298 among the 1000 genomes populations. The allele frequency of the reference and alternative allele is visualized by the geography of genetic variants browser developed by the university of Chicago. C) P-value of the single variant genome-wide association test for the sixty-four laboratory assessments at the lead SNP rs6020298. The P-value of the three traits (Severity, Severity score and Disease Duration) in Figure 3 were also displayed.

**Figure 5.**
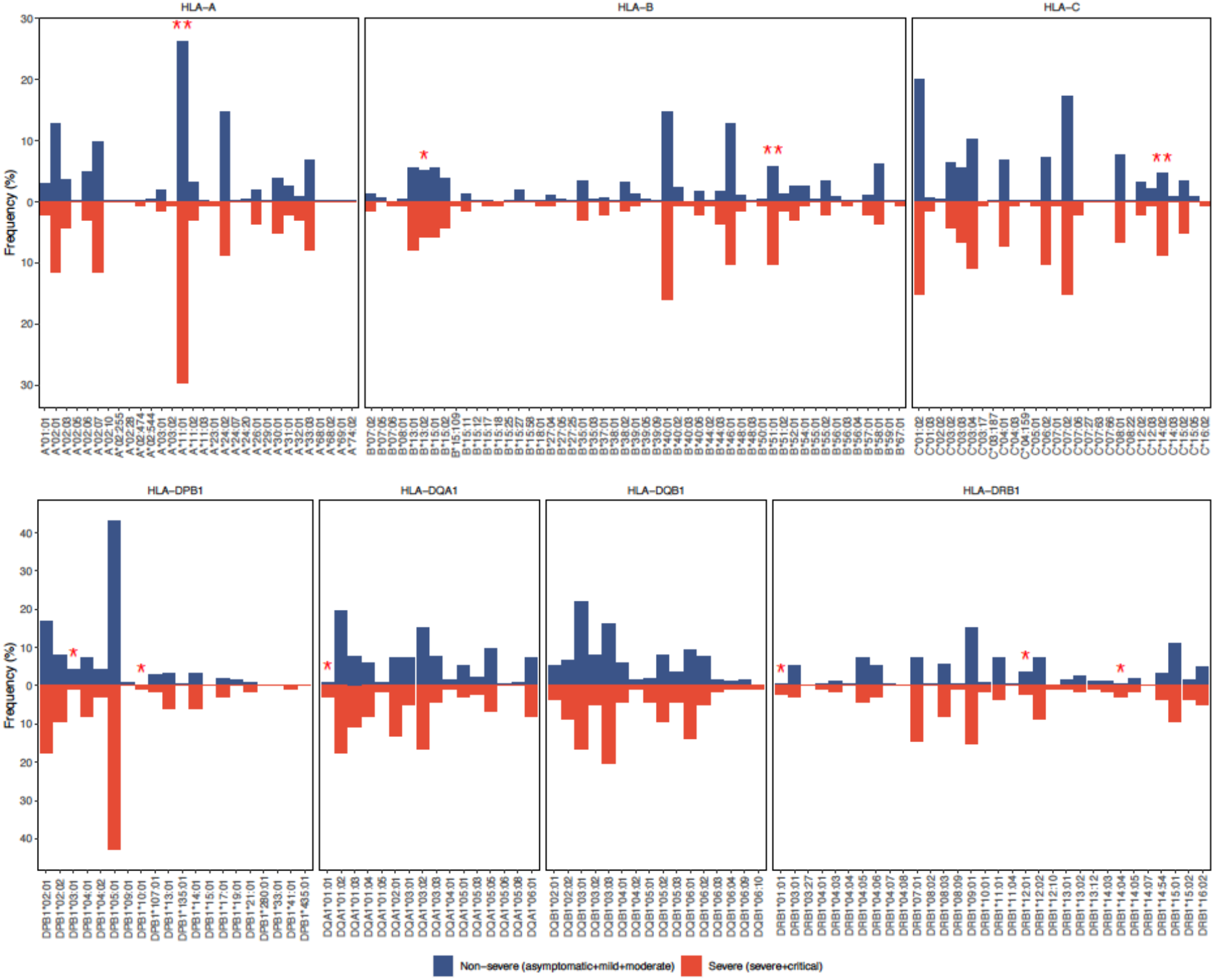
Human leukocyte haplotype allele frequency between severe vs non-severe groups. Comparison for class I HLA genes (top). Comparison for class II HLA genes (bottom). Star indicates significance level in a logistic regression on the allele frequency with age, gender and the top twenty principal components as covariates. *<0.05, **<0.01

We further performed optimal SKAT gene-based association test on the functional variants including a total of 99,166 missense and loss of function variants that were predicted to have high or moderate impacts by variant effect predictor among the patients. The *NOA1* gene tend to higher mutation burden in the severe group (P= 8.1e-07) (**Figure 3D**). This gene encodes the GTPase that functions in the mitochondrion and has been associated with platelet count and leukocyte count^45^. We didn’t identify other genes that are genome-wide significantly associated with the severity score or the disease duration (**Figure3 E-F**).

### HLA gene alleles associated with severity in the COVID-19 patients

Manifestation of numerous infectious diseases are closely related to the genetic variants across the major histocompatibility complex (MHC) genes, i.e. the human leukocyte antigen (HLA) genes, which play an essential role in presenting the antigen determinant epitopes from the pathogens to the T cell or B cell to activate the host immune response^50,51^. In the 2003 severe acute respiratory (SARS) outbreak, caused by the SARS coronavirus (SARS-CoV) related to SARS-CoV-2, the HLA-B*46:01 was reported to be associated with infection severity in East Asian patients^25^. Herein, we investigated the genetic effect from HLA genes on the COVID-19 patient severity. We re-aligned all the reads mapped to the eight HLA haplotypes in the human reference genome (GRCh38) and all the unaligned reads and typed the three class I HLA genes (A, B, C) and four class II HLA genes (DPB1, DQA1, DQB1, DRB1) using the xHLA^52^ and the SOAP-HLA approach^53^. 4-digit haplotyping resolution was achieved for 99% of the patients for all the genes except for DQA1 where three patients were only typed to the 2-digit resolution. We observed zero mendelian error rate for the typing results using the family members involved in the study. We investigated whether some HLA alleles may significantly differ between the broadly defined severe (severe and critical, N=69) and mild (asymptomatic, mild and moderate, N=215) groups of unrelated patients using a logistic regression with age, gender and the top 20 principal components as covariates. The frequency comparison between the severe and mild groups for the total 30 HLA-A, 51 HLA-B, 28 HLA-C, 20 DPB1, 21 DQA1, 16 DQB1 and 32 DRB1 alleles were displayed in **Figure 4 and Table S2**. Among the class I HLA genes, C*14:02 (severe 8.7% vs mild 4.6%, OR=4.7, P=3e-3), B*51:01 (severe 10.1% vs mild 5.8%, OR=3.3, P=7e-3), A*11:01 (severe 29.7% vs 26.2%, OR=2.3, P=8.5e-3) are the top three most significant alleles between the two groups that predispose the patients entering the severe stage **(Table 1**). The HLA-A*11:01, B*51:01 and C*14:02 is in strong linkage equilibrium with each other and thus represents one haplotype. This haplotype has an average allele frequency 2.4% - 3.6% among the Chinese populations according to the HLA Allele Frequency Net Database^54^. In our study, we find that this haplotype is more prevalent in the severe patients compared with the mild patients.

**Table 1.**
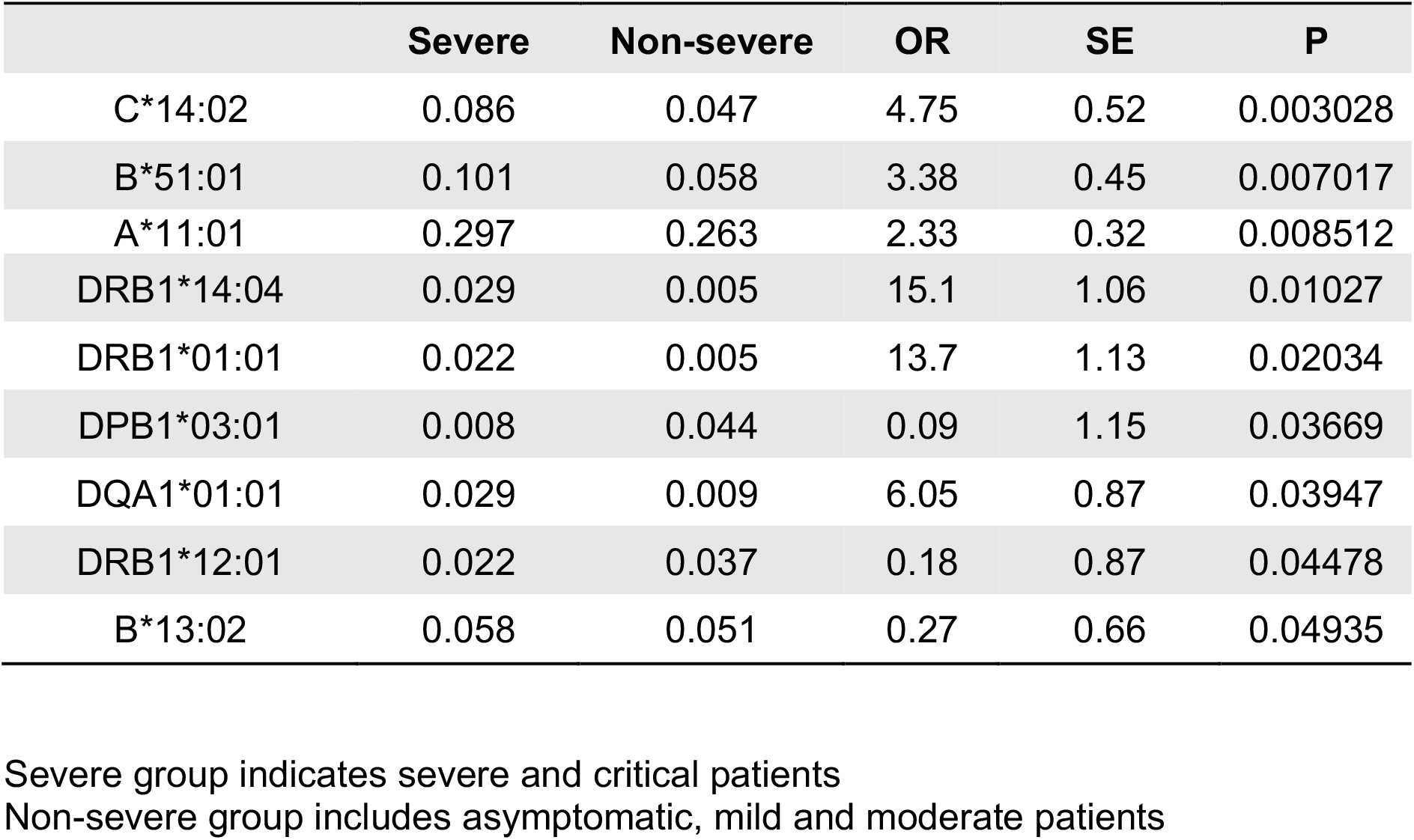
Nominal association of HLA allele and severity by logistic regression.

|  | Severe | Non-severe | OR | SE | P |
| --- | --- | --- | --- | --- | --- |
| C*14:02 | 0.086 | 0.047 | 4.75 | 0.52 | 0.003028 |
| B*51:01 | 0.101 | 0.058 | 3.38 | 0.45 | 0.007017 |
| A*11:01 | 0.297 | 0.263 | 2.33 | 0.32 | 0.008512 |
| DRB1*14:04 | 0.029 | 0.005 | 15.1 | 1.06 | 0.01027 |
| DRB1*01:01 | 0.022 | 0.005 | 13.7 | 1.13 | 0.02034 |
| DPB1*03:01 | 0.008 | 0.044 | 0.09 | 1.15 | 0.03669 |
| DQA1*01:01 | 0.029 | 0.009 | 6.05 | 0.87 | 0.03947 |
| DRB1*12:01 | 0.022 | 0.037 | 0.18 | 0.87 | 0.04478 |
| B*13:02 | 0.058 | 0.051 | 0.27 | 0.66 | 0.04935 |

Severe group indicates severe and critical patients
Non-severe group includes asymptomatic, mild and moderate patients

Notably, although B*46:01 has been suggested to present the fewest SARS-CoV and SARS-CoV-2 peptides in an in silico analysis^55^ and has been associated with the SARS-CoV in a small sample size association analysis without correcting demographic and geographic covariates^25^, our analysis doesn’t support this allele is associated with the disease severity (OR=0.5, P=0.15). On the contrary, allele frequency of B*46:01 is less frequent in the severe patients (10.1%) than among the mild patients (12.8%). Class II HLA genes is less significantly associated with the disease severity compared to the Class I genes (**Table 1**). DRB1*14:04 (severe 2% vs mild 0.5%, P=0.01), DRB1*01:01 (severe 2.2% vs 0.5%), DQA1*01:01 (severe 2.9% vs 0.9%) are the top three risk alleles while DPB1*03:01 (severe 0.7% vs mild 4.5%) and DRB1*12:01 (severe 2.2% vs mild 3.7%) might display a protective effect.

### Comparison with general population for potential genetic contribution to SARS-CoV-2 infection susceptibility

Our study till now has been restricted in the infected patients to understand genetic contribution to patient severity. Mapping genes related to infection susceptibility is more difficult. The ideal design commands a comparison between people who are exposed or not exposed to the pathogen. This is hard to meet because early detection and isolation of infected patients are the primary containment strategies against an outbreak^56^. Therefore, we choose another approach to investigate genetic susceptibility by comparing the 284 unrelated hospitalized patients (the Case) with two general populations including 301 Chinese individuals in 1000 genome project^36^ (the Control I) and 665 individuals recruited from the Chinese Reference Panel program (CNPR, manuscript in preparation, the Control II). Control I and Control II differ in terms of the similarity of the adopted sequencing protocol compared to the Case. All the technical components are almost the same between the Case and Control 2 except for sequencing depth (case 46x versus control 2 30x).

On the other hand, various factors are different between the Case and Control 1, including types of sample (case fresh blood versus control 1 cell line), sequencing technology (case MGI’s nanoball sequencing versus control 1 Illumina sequencing), sequencing read cycles (case 100bp pair-end versus control 1 150bp pair-end) and the sequencing depth (case average 46x versus average 7x). Study like this can reveal genetic difference between the infected population and the general population if any and if not, instruct on what cautions should be taken when comparing the disease cohorts versus the general in the whole genome sequencing context.

We analyzed the data carefully by jointly genotype the samples from their individual gvcf files using the GATK best practices^39^ instead of simply merging the population vcf files of the case and the control. Principle component analysis indicates that population structure is the dominant confounding factor and sequencing induced batch effects were difficult to identify in the PCs (**Figure S17, Figure S18**). Similarly, we conducted both single variant and gene-based association tests for the two case-control data sets using the top 20 PCs, gender and age (age was not available for 1KGP samples and was used for the CNPR alone) as covariates. Surprisingly, in the single association test for the high and moderate impact variants, many variants in the HLA region displayed significant associations between the COVID-19 patients and the 1KGP Chinese (**Figure 6A**) even though the inflation was seemingly adequately controlled (**Figure S19)**. In the gene-based association test, we observed significantly different mutation burdens in the immunoglobulin loci (**Figure 6B**). However, this was not replicated when we compared the COVID-19 patients with the 665 CNRP individuals (**Figure 6C-D**). Therefore, we inferred that the association signals between the 1KGP and the COVID-19 patients were probably due to sequencing batch effects. As the fresh blood of an infected individual contains numerous somatic mutated B-cells, patients tend to accumulate more mutations in the immunoglobulin genes^57^. As many studies try to directly compare the allele frequency between the general population and the COVID-19 patients^30,32^, our discoveries remind us of the necessity for re-evaluation of the significant hits given distinct experimental protocol for case and control.

**Figure 6.**
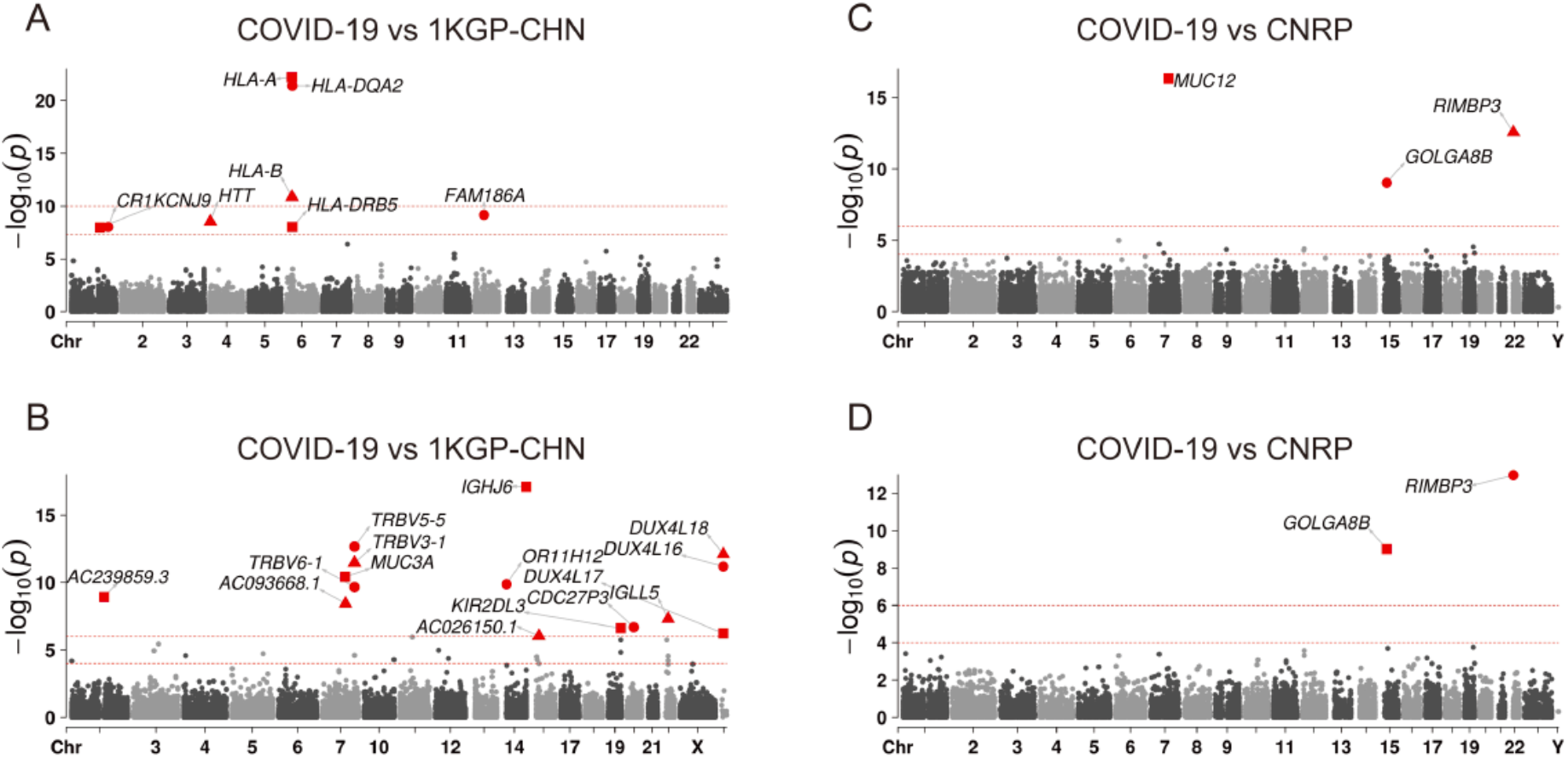
Single variant and gene-based association test between COVID-19 patients and the general populations. (A) single variant association test and (B) gene-based association test between the unrelated COVID-19 patients (N=284) and the 1KGP Chinese population (N=301) (C) single variant association test and (D) gene-based association test between the unrelated COVID-19 patients (N=284) and the CNRP Chinese population (N=665). Only variants with moderate or high impacts by variant effect predictor were shown in (A) and (C).

In the single variant association test between the COVID-19 patients and the CNPR who were sequenced using the same experimental protocol and were laboratory PCR tested negative, we identified genome-wide significant associated signals tagged by a novel missense variant (Patient T allele frequency=0.34, CNPR T_AF=0.14, OR=18, P=4,7e-17) in *MUC2*; a missense variant rs200584390 (Patient G allele frequency=0.31, CNPR G_AF=0.09, OR=9.29, P=1.5e-13) in *RIMBP3* and a missense variant rs200975425 (Patient T allele frequency=0.24, CNPR T_AF=0.39, OR=5.4, P=9.4e-10) in *GOLGA8B* (**Figure 6C**). Gene-based association test also indicates that *RIMBP3* and *GOLGA8B* were different between the patients and the CNPR (**Figure 6D**). Those discoveries require further replication and interpretation when more sequencing data for patients and for general populations become available worldwide^33^.

## Discussion

We have conducted the first genetic association study for the COVID-19 severity and SARS-CoV-2 infection susceptibility by studying the genome and clinical outcome of 332 patients in a designated infectious disease hospital in the Shenzhen City. Instead of using the microarray or the exome genome sequencing, we have carried out high-depth whole genome sequencing and analysis for the patients to obtain the greatest possible power given a small sample size available so far. The study design enables the detection of very rare and private functional variants for the patients^58^ and ensures that the potential causal variants are directly assayed to compensate the loss of power due to poor linkage disequilibrium between the assayed and the causal variants^59^.

We revealed that the disease progression after the SARS-CoV-2 infection was a complex event and not explained by a monogenic model. The severe and critical patients did not carry causal monogenic variants related to the disease severity in their genome. We identified that the missense variant rs12329760 in *TMPRSS2* was less frequent among the critical patients compared to the rest of the patients and the general population. This variant results in an alteration of the valine to the methionine at the 197th amino acids (p.Val197Met) has been predicted to decrease the TMPRSS2 protein stability and ACE2 binding^44^. On the other hand, our study using Chinese samples did not support the assumption^30^ that host genetic factors in the essential SARS-CoV receptor *ACE2* and some other genes involved in the host pathogen interaction network might play a role in determining the patient’s severity or susceptibility.

In the genome-wide association analysis, a gene locus around *TMEM189-UBE2V1* displayed suggestively association with COVID-19 severity. This gene locus contains genes such as *UBE2V1* and *TEMEM189-UBE2V1* that are known to function in the interleukin-1 signaling pathway^45,46^. The lead SNP rs60220284 is an eQTL where the risk allele A increases the gene expression of genes within the locus^60^ and is more prevalent in the severe and critical patients. While COVID-19 severe patients demonstrate elevated IL-1 compared to the mild patients and the general population^47^, our study suggests potential correlation between genetic variability in this gene and the disease severity.

Notably, the HLA-A*11:01, B*51:01 and C*14:02 alleles were significantly more prevalent in the severe and critical severe patients compared to the mild and the moderate patients after careful control of population structure and demographic characters such as age and gender. The three alleles were in linkage disequilibrium with each other and has been previously reported to have a 2-3% population allele frequency in Dai and Jinpo minorities in China^54^ and the B*51:01 has been previously linked to the Behcet’s disease^61^, a kind of rheumatic disease. We were not able to access the role of HLA-B*46:01, although it has been predicted as the worst presenting HLA alleles to the SARS-CoV-2 proteome^55^ and linked to the SARS 2003 outbreak^25^.

Surprisingly, genome-wide association study using the COVID-19 patients as the case and the 1000 genome Chinese population as the control suggested an enrichment of significantly associated signals in the HLA region and mutation burden in the immunoglobulin genes. Nonetheless, this was not replicated when we compared the patients to another independent Chinese population. A lot of efforts in the genetic field have been made and there may be more in the future to investigate genetic susceptibility of the SARS-COV-2 infection by directly comparing two or more general populations with the COVID-19 patients^32,33^. Therefore, cautions should be taken to properly control the batch effects. Replication is essential and perhaps a joint-analysis effort can rule out the real signals from the false delusion.

Some limitations of the study should be noted. Power analysis indicates that sample size of around 300 is barely sufficient to identify genome-wide significant genetic variants with minor allele frequency greater than 0.2 and odds ratio greater than 1.8 given type I error rate 0.05. We don’t have power to detect causal variants beyond this risk and allele frequency scenario. In addition, although the study of hospitalized patients in a designated hospital includes all severe patients, the design has a limited presentation of the asymptomatic patients (7.5%) which ratio has been estimated to be 30.8% (95% confidence interval 7.7-53.8%)^62^. Given that RT-PCR test and the seroprevalence immunoglobulin M and G antibody tests targeting the SARS-CoV-2 has been widely adopted in China and around the globe, it will be important to identify and study the extreme asymptomatic patients to understand the host factors contributing to a capable control of the viral infection.

As we and the others are continuing to recruit patients and data in China and around the world to understand the host genetic background underlying the varying clinical outcome of the patients, this work represents the first genetic study on the Chinese hospitalized patients where high quality sequencing data were generated and systematic analysis on the genomic and clinical data were conducted. Our results highlight several genetic factors involved in the immune responses including genes involved in the viral entry in the host cells, genes related to immune responses and the HLA alleles. This work is also an important and initial start to guide study design regarding the selection of samples, the genetic assay approach, the bioinformatics and the statistical genetic analysis for COVID-19 as well as other infection and complex disease. The publicly available summary statistics will encourage international collaborative efforts to understand the host-pathogen interaction and to contain the COVID-19 outbreak.

## Data Availability

The data that support the findings of this study, including the allele frequency for the five groups of patients at all the 20 million detected genetic variants and the genome-wide association test summary statistics have been deposited in CNSA (China National Genebank Sequence Archive)in Shenzhen, China with accession number CNP0001107 (https://db.cngb.org/cnsa/).

## Acknowledgements

The study was supported by National Natural Science Foundation of China (31900487), Guangdong Provincial Key Laboratory of Genome Read and Write (No. 2017B030301011) and China National GeneBank (CNGB). We would like to acknowledge Fan Zhang from Illumina, Zilong Li and Kang Fang, Defu Xiao from BGI, Xinjun Zhang from University of California, Los Angeles, Emilia Huerta-Sanchez from Brown University and Rasmus Nielsen from University of UC Berkeley for helpful discussions of the results and advice.

## Author contributions

Conceptualization, L.L, X.J., Q. H., S. Liu, S.H.; Methodology, J.S. S. Liu, Y.Z., X.T.;Formal Analysis, S.H., Y.Z., X.Q., Zhi.L., P.L., Y.H., R.L., X.T., Y.B., S. Liu; Resources, F.W., R.G., C.L,W.X., Zhi.L, Q.T. R.C. X.L, X.Z., G.D.; Data Curation, S. Liu, F.W., R.G., C.L; Writing – Original Draft, S. Liu; Writing – Review & Editing, All; Supervision, X.X.,J.W.,H.Y.; Project Administration, X.J., S.H., S. Liu, and F.C.; Funding Acquisition, F.W., X.J. and S. Liu.

## Material and Methods

### Patient recruitment and definition of phenotypes

A total of 332 patients were recruited from Jan 11th 2020 to Apr 2020 in Shenzhen Third People’s Hospital, the only referral hospital in Shenzhen City, China^34^. All were confirmed with SARS-COV-2 infection using real-time reverse-transcriptase– polymerase-chain-reaction (RT-PCR) assay of nasal and pharyngeal swab specimens. The demographic, epidemiological, clinical and laboratory assessments were extracted from the electronic medical records of the patients. This study was approved by the ethics commissions of the Shenzhen Third People’s Hospital Ethics Committee with a waiver of informed consent. According to the 5th edition of the national treatment guideline of COVID19 in China and the Chinese CDC criteria^6^, the patients were diagnosed as asymptomatic, mild, moderate, severe and critically severe according to the most severe stage they experienced during the disease course. The asymptomatic, mild and the moderate groups of patients do not experience pneumonia. When meeting any one of the following criteria, 1) RR>30 2) Oxygen level < 93% 3) PaO2/FiO2 < 300 mmHg 4) disease progression greater than 50% area in CT scan, a patient is categorized as severe patients. Patients experienced one of the following 1) respiratory failure and requires mechanical ventilation 2) shock 3) complicated by failure of other organs and requires intensive care monitoring were classified as critically severe.

### Assignment of severity score to each patient

A machine learning XGBoost-based model was developed to predict ordinal severity scores using patients’ phenotype data of 64 laboratory test results^63^. We first filtered out the laboratory test items of which at least 50% of patients did not have any recordings. The remaining 52 laboratory test items with missing values were further imputed by missForest algorithm^64^. The missForest is a nonparametric method to impute missing values using random forest model in an iterative fashion. Then the originally ordered severity levels of asymptomatic, mild, moderate, severe and critical were assigned integer values of 1, 2, 3, 4 and 5, respectively. The numeric representations retained the ordinal levels of severity. We applied the reduction framework mentioned in Li et al^65^, where the ordinal regression was reduced to binary classification. The reduction framework of extended binary classification was then integrated within XGBoost model. Moreover, we selected the most predictive laboratory test items using SHAP (SHapley Additive exPlanations) algorithm^66^. The SHAP is a game theoretic approach to explain the output of a given machine learning model using Shapley values from game theory and their related extensions. We finally trained the XGBoost-based ordinal regression model using the selected laboratory test items. As a result, the prediction outcome produced by the final model was typically a real number reflecting severity level that was used in the downstream analysis. We used 100 base estimators for missForest, maximum iteration of 10, and the criterion was mean squared error. For the XGBoost-based ordinal regression model, we used 500 base estimators and learn rate of 0.5. In general, the hyper-parameters of models in this study were chosen by combining grid search of 5-fold cross validation and manual tuning.

### DNA extraction, library construction and deep whole genome sequencing

Genomic DNA was extracted from frozen blood samples of the 332 patients using Magnetic Beads Blood Genomic DNA Extraction Kit (MGI, Shenzhen, China). At least 0.5μg was obtained for each individual and used to create WGS library, which insert sizes 300-500bp for paired-end libraries according to the BGI library preparation pipeline. Sequencing was conducted on the DNBSEQ platform (MGI, Shenzhen, China) to generate 100bp paired-end reads.

### Genome alignment and variant detection

We used Sentieon Genomics software (version: sentieon-genomics-201911) to perform genome alignment and variant detection^67^. Analysis pipeline were built according to the recommendation in the Broad institute best practices described in https://gatk.broadinstitute.org/hc/en-us/sections/360007226651-Best-Practices-Workflows. Sequencing reads were mapped to hg38 reference genome using BWA algorithm. For each sample, after remove duplicates, Indel realignment and base quality score recalibration (BQSR), SNP and short Indel variants were detect using the Sentieon Haplotyper algorithm with option -- emit_mode gvcf to generate an individual GVCF file. Then the GVCF files for all samples were subjected to Sentieon GVCFtyper algorithm to perform joint variant calling.

### Variant Quality Score Recalibration and Filtration

Variant Quality Score Recalibration were perform using Genome Analysis Toolkit (GATK version 4.1.2). Known variant files were downloaded from the GATK bundle. For indel recalibration, we used Mills_and_1000G_gold_standard indels as the positive training and true set. For SNP recalibration, we used hapmap_3.3, 1000G_omni2.5, and 1000G_phase1.snps as positive training sets, hapmap_3.3 as true set, and dbSNP_v146 as the known set. The metrics DP, QD, MQRankSum, ReadPosRankSum, FS, SOR were used in the recalibration process. The truth-sensitivity-filter-level were set to 99.0 for both the SNPs and the Indels. Finally, variants with quality score >= 100 were selected for further analysis.

### Familial relationship and population structure analysis

PLINK (v1.9)^68^ and KING (v2.1.5)^69^ was applied to detect the kinship relatedness between each pair of the individuals. 48 patients from 16 families were detected as related to each other. For several allele frequency-based approach, we exclude the related patients and thus the sample size was restricted to 284. PCA was performed using a subset of autosomal bi-allelic SNPs on the unrelated patients using PLINK (v1.9). The PC-AiR module (Principal components analysis in related samples) in the Genesis R package was used to conduct PCA analysis for the 332 patients including the related family members. Several restrictions were employed to select the final 614,963 SNPs for PCA analysis, including minor allele frequency (MAF) ≥ 1% (common and low-frequency variants), genotyping rate ≥ 90%, Hardy-Weinberg-Equilibrium (HWE) P > 0.000001, and removing one SNP from each pair with r2 ≥ 0.5 (in windows of 50 SNPs with steps of 5 SNPs).

### Genotype-phenotype association analysis

We have applied both the rvtest^70^ and the SAIGE^71^ approaches to carry out logistic regression, linear regression, burden test, the sequence kernel analysis test (SKAT) and the optimal SKAT-O algorithm for the genotype-phenotype association tests using the default parameters. For all the association tests, we have used the gender, the age and the top 20 principal components from the principal component analysis as the covariates. Exception is for the GWAS between the 1KGP and the COVID-19 patients as age is not available for the 1KGP data set. Independent loci were defined as significant variants clustered in a 1Mbp window. The lead SNP was defined as the SNP in the 1Mbp window that has most significant, i.e., smallest p value. The genomic inflation factor, GC lambda, attenuation ratio, LD score regression intercept and the SNP heritability were estimated using the LD score regression approach ^72^. The qqman R package was applied to generate the manhattan and qqplot. We defined genome-wide significance for single variant association test as 5e-8, suggestive significance as 1e-5 and for gene-based association test as 1e-6.

### HLA typing

When performing HLA typing, we first extracted reads which aligned to HLA region of GRCh38 and unmapped reads from individual bam files. Then using xHLA algorithm^23^ typing HLA class I(A B C gene) and II(DRB1 DQB1 DPB1) genes. DQA1 gene was typed using SOAP-HLA algorithm^53^ for xHLA does not include this gene. We performed the association analysis between HLA types and the binary severe and mild groups using PLINK (version 1.90) using a logistic regression model, adjusted for age, gender and top 20 PCs.

